# Environmental scan of provincial and territorial planning for COVID-19 vaccination programs

**DOI:** 10.1101/2020.12.22.20248685

**Authors:** Shannon E. MacDonald, Hannah Sell, Sarah Wilson, Samantha B. Meyer, Arnaud Gagneur, Ali Assi, Manish Sadarangani, And members of the COVImm study team

## Abstract

**Background:** Public health departments in Canada are currently facing the challenging task of planning and implementing COVID-19 vaccination programs.

**Objective:** To collect and synthesize information regarding COVID-19 vaccination programs in each of the provinces and territories (P/Ts).

**Methods:** Provincial/territorial public health leaders were interviewed via teleconference between August-October 2020 to collect information on the following topics, drawn from scientific literature and media: unique factors for COVID-19 vaccination, adoption of National Advisory Committee on Immunization (NACI) recommendations, priority groups for early vaccination, and vaccine safety and effectiveness monitoring. Data were grouped according to common responses and descriptive analysis was performed.

**Results:** Eighteen interviews occurred with 25 participants from 11 of 13 P/Ts. Factors unique to COVID-19 vaccination included prioritizing groups for early vaccination (*n*=7), public perception of vaccines (*n*=6), and differing eligibility criteria (*n*=5). Almost all P/Ts (*n*=10) reported reliance on NACI recommendations. Long-term care residents (*n*=10) and health care workers (*n*=10) were most frequently prioritized for early vaccination, followed by people with chronic medical conditions (*n*=9) and seniors (*n*=8). Most P/Ts (*n*=9) are planning routine adverse event monitoring to assess vaccine safety. Evaluation of effectiveness was anticipated to occur within public health departments (*n*=3), by researchers (*n*=3), or based on national guidance (*n*=4).

**Conclusion:** Plans for COVID-19 vaccination programs in the P/Ts exhibit some similarities and are largely consistent with NACI guidelines, with some discrepancies. Further research is needed to evaluate the success of COVID-19 vaccination programs once implemented.

## Introduction

The race for the development of COVID-19 vaccines is well underway, with the first vaccines now approved for use in Canada (1). Canadian public health officials are now facing the next major challenge of the pandemic: planning and implementing vaccination programs. There are many decisions that need to be made, including the distribution, administration, and monitoring of vaccines (2). As initial vaccine supply is limited, one important consideration is the prioritization of target groups for COVID-19 vaccination. The National Advisory Committee on Immunization (NACI) have released guidance outlining key populations for receiving initial vaccine supply (3,4). However, it is ultimately up to each of the provinces and territories (P/Ts) whether to follow these guidelines, and to determine the logistics of administering and monitoring COVID-19 vaccination programs. The objective of this study was to collect and synthesize information regarding planned COVID-19 vaccination programs in each of the P/Ts, including logistic considerations, priority groups, and vaccine safety and effectiveness monitoring.

## Methods

This pan-Canadian environmental scan involved key informant interviews of public health leaders from P/Ts across Canada. Participants were recruited via convenience sampling through referrals from members of the research team, P/T Ministries of Health, and the NACI Secretariat at the Public Health Agency of Canada. These key informants were contacted via an initial email sent by the NACI Secretariat, inviting them to participate in the current study. Interested individuals were emailed an information sheet and consent form. To optimize response rate, up to two email reminders were sent. Some participants were identified through snowball sampling, with study participants suggesting additional key informants. Interviews took place from August to October 2020, prior to release of NACI’s preliminary guidance (3). Interviews (35-60 minutes long) were conducted by members of the research team (HS, AA, MK).

Interview questions included key topics related to COVID-19 vaccination, as identified in scientific literature and news articles, and augmented with input from the research team and knowledge users, including the NACI Secretariat. The interview guide consisted of mainly open-ended questions about the following topics: unique factors to be considered in COVID-19 vaccination program planning, the extent of reliance on NACI recommendations, the use of a geographical prioritization framework for vaccine allocation, target groups for prioritization for early vaccination, and plans for monitoring vaccine safety and effectiveness. The interview guide was pilot tested to check face and content validity, flow, and comprehension. Ethical approval for this study was obtained from the Health Research Ethics Board at the University of Alberta.

Interviews were transcribed verbatim and any personally identifying information was removed. Interview data were coded and grouped according to common responses and then quantified. Descriptive analysis of response counts was performed using Microsoft Excel.

## Results

Invitation emails from NACI were sent to 35 potential participants: 13 agreed to participate, 1 declined, and 21 did not respond. Five participants were recruited via referrals from other participants. Prior to the interview, participants were told they could invite other colleagues to provide additional perspectives. Therefore, some interviews contained more than one participant. In total, there were 18 interviews with 25 participants from 11 of the 13 P/Ts.

Twelve participants provided a P/T-level perspective, 9 provided a regional/municipal perspective, and 4 provided a combination of P/T and regional/municipal perspectives. Common job titles of participants included Director of Immunization or Communicable Disease Control (*n*=2), Immunization Program or Policy Manager (*n*=7), Medical Officer of Health (*n*=5), Public Health or Medical Consultant (*n*=3), Policy Analyst (*n*=2), Public Health or Communicable Disease Specialist (*n*=2), and Other (*n*=4). Participant responses were synthesized and presented by P/T.

### Unique factors for COVID-19 vaccination programs

A wide array of factors that are unique to planning for COVID-19 vaccination programs were identified (see Table 1). Slightly over half of P/Ts (*n*=7) indicated the need to prioritize target groups for early vaccination. Many P/Ts (*n*=5) also highlighted the possibility of having different eligibility criteria for each vaccine (i.e., if one vaccine is more effective in older adults), which may impact the order of priority groups.

**Table 1.** Unique factors to consider for COVID-19 vaccination programs, by number of provinces/territories (N=11)

| Unique factor | Number of P/Ts,<sup>a</sup> *n* |
| --- | --- |
| **Priority groups** |  |
| Prioritization of target groups | 7 |
| Differing eligibility criteria | 5 |
| Equity in delivery | 1 |
| **Public engagement** |  |
| Public perception of the vaccine, including vaccine hesitancy | 6 |
| Clear communication with public | 3 |
| **Logistics and supply** |  |
| Logistics, storage, cold-chain management | 4 |
| Limited vaccine supply, availability of vaccine | 3 |
| Availability of PPE and other vaccination supplies (other than the vaccine itself) | 3 |
| Vaccine distribution | 2 |
| Resource issues (in general) | 2 |
| Vaccine procurement | 1 |
| **Delivery** |  |
| COVID-19 related restrictions, public health measures, PPE | 4 |
| Vaccine provider (e.g., physicians, pharmacists, public health) | 4 |
| Appointment-based delivery vs. mass clinics | 3 |
| Need to vaccinate everyone, large volume of people | 3 |
| Less human resources due to COVID-19 redeployment | 3 |
| Training for providers | 2 |
| Uncertainty, not having enough information to plan | 2 |
| **Vaccine characteristics** |  |
| Possibility of needing more than one dose | 4 |
| Vaccine safety | 3 |
| Dealing with a new vaccine | 3 |
| Considerations for the route of administration | 3 |
| Possibility of having more than one vaccine | 2 |
| Speed with which vaccine development is occurring | 2 |

Note. P/T = province/territory; PPE = personal protective equipment
<sup>a</sup>Some P/T responses fell into more than one category

Some P/Ts also discussed factors related to public engagement, including having clear communication with the public regarding safety implications, eligibility criteria, and priority groups (*n*=3). Likewise, six P/Ts highlighted the need to manage public perception of COVID-19 vaccines. Specifically, one P/T felt that vaccine hesitancy for COVID-19 vaccines would be greater than for previous vaccines.

P/Ts also discussed unique factors related to logistics and supply of COVID-19 vaccines. Four P/Ts highlighted the unique storage requirements of some of the vaccines, with some P/Ts stating that it was unlikely that all providers currently had the capacity to store vaccines at the appropriate temperature. Others noted that supply of the vaccine (*n*=3) and other vaccination supplies (*n*=3) will likely be limited.

Planning for the delivery of the COVID-19 vaccines was anticipated to be challenging, with some P/Ts (*n*=4) reporting that they were unsure about which providers would deliver the vaccines (e.g., public health, physicians, pharmacists), or whether they would have appointment-based clinics or mass clinics (*n*=3). Similarly, four P/Ts mentioned the need for adapting vaccination clinics to follow COVID-19 restrictions, including physical distancing, personal protective equipment, layout, one-way flow of traffic, and ventilation. One P/T mentioned including industrial engineers on their planning team to consider these factors. A full list of P/T responses is provided in Table 1 below.

### Reliance on NACI recommendations

Almost all P/Ts (*n*=10) indicated that they would likely rely on the NACI recommendations for target groups in planning their COVID-19 vaccination strategies. One P/T indicated that they would more likely rely on their provincial/territorial immunization committee recommendations.

### Use of a geographical prioritization framework

None of the P/Ts had firm plans for a geographical prioritization framework based on disease incidence (i.e., target groups in high COVID-19 incidence areas are prioritized over target groups in low incidence areas). The majority of P/Ts (*n*=7) were open to this approach if advised by NACI (*n*=1), or if the vaccine characteristics (*n*=1) or number of doses available (*n*=3) warrants it. Three P/Ts were against using a geographical prioritization framework due to concerns with the equity of this approach (*n*=1) or due to their jurisdiction’s small geography or dense population (*n*=2). One P/T did not know if they were planning on using a geographical prioritization framework.

### Priority group ranking

As shown in Figure 1, participants were asked to rank their top 5 priority groups, with rank 1 representing the group that should receive COVID-19 vaccination first. For reporting purposes, we used the ranking of the respondent from each P/T that had the most cross-provincial perspective. One P/T did not provide a ranking, for a total of 10 P/Ts. All of the P/Ts ranked long-term care residents (*n*=10) and health care workers (*n*=10) in the top 5 priority groups for receiving COVID-19 vaccination. Most P/Ts also ranked people with chronic medical conditions (*n*=9) and seniors (*n*=8), followed by people of Indigenous ancestry (*n*=4), those with socioeconomic disadvantage (*n*=3), infants/children (*n*=2), people living in remote communities (*n*=2), and new immigrants and refugees (*n*=1).

**Figure 1:**
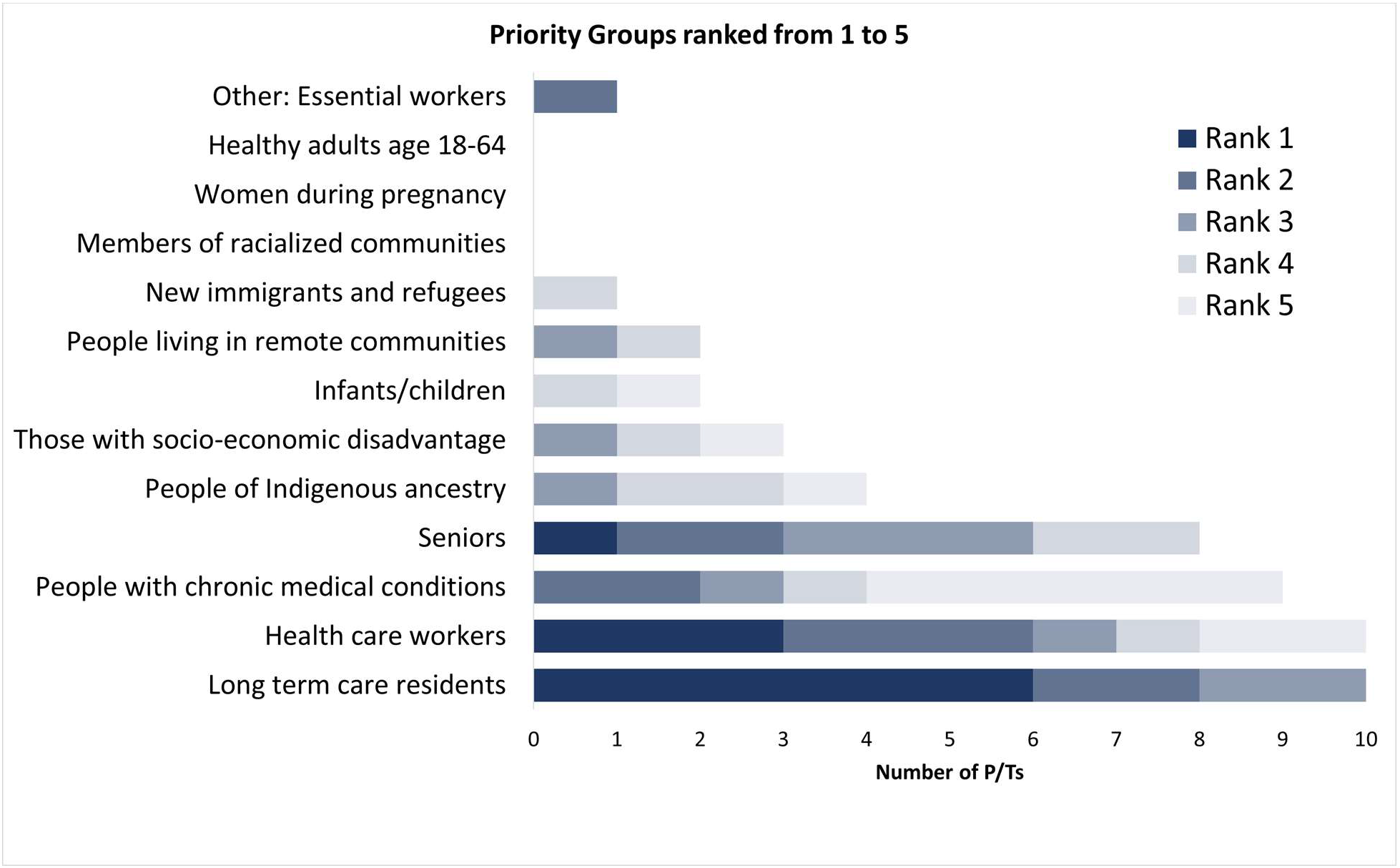
The number of provinces/territories that chose each group in their top 5 groups to be prioritized for early vaccination in the presence of limited vaccine supply (N=10)^a,b^. **Footnotes:** ^a^One province/territory chose not to answer this question ^b^For those who selected seniors (*n*=8), 7 indicated that they would target seniors aged 65+ years, while 1 indicated that they would target those 60+ years.

### Monitoring vaccine safety and effectiveness

In regards to post-market vaccine safety monitoring, most P/Ts (*n*=9) planned to conduct their routine adverse event monitoring, while some (*n*=3) anticipated enhanced surveillance of adverse events (see Table 2). Some P/Ts (*n*=4) anticipated that this will be done by federal/provincial/territorial committees and groups. For post-market vaccine effectiveness monitoring, some P/Ts (*n*=3) anticipated that their P/T public health departments would do this, with a similar number (*n*=3) stating that this will be routine information collected. Others (*n*=3) expected that this will be done by researchers or research organizations.

**Table 2.** The number of provinces/territories that indicated each approach to COVID-19 vaccine safety and effectiveness monitoring (N=11)

| Planned approach | Number of P/Ts, $n^a$ |
| --- | --- |
| **Safety** |  |
| Regular adverse event reporting | 9 |
| Enhanced surveillance of adverse events | 3 |
| Reliance on federal/provincial/territorial committees and groups (e.g., CIC, CIRC) | 4 |
| Reliance on what NACI recommends | 1 |
| Undecided | 1 |
| Don't know | 2 |
| **Effectiveness** |  |
| Reliance on provincial public health (e.g., surveillance teams) | 3 |
| Reliance on researchers/research organizations | 3 |
| Reliance on NACI or other national guidance | 4 |
| Collect routine monitoring information (e.g., number of clients who tested positive after vaccination, vaccine coverage) | 3 |
| Undecided | 2 |
| Don't know | 1 |
| No answer | 2 |

Note. CIC = Canadian Immunization Committee; CIRC = Canadian Immunization Registries and Coverage Network; NACI = National Advisory Committee on Immunization; P/T = province/territory
<sup>a</sup>Some responses by P/T fall into more than one of the above categories

## Discussion

Although P/T rankings of potential priority groups were collected prior to the publication of NACI’s guidance documents, the overall P/T rankings aligned somewhat with NACI recommendations. Specifically, the groups ranked highest in this study were health care workers, long-term care residents, followed by people with chronic medical conditions, and seniors. The most recent NACI recommendations prioritize health care workers, long-term care residents and staff, seniors aged 70 and older (with those 80+ years having highest priority), and adults in Indigenous communities (4). The notable difference between the P/T ranking in our study and NACI recommendations, is that less than half of P/Ts ranked Indigenous communities in the top five prioritized groups, and people with chronic medical conditions (ranked third by most P/Ts) were not included in NACI’s most recent guidance on early vaccination (4).

A common consideration among P/Ts was the potential negative public perception of COVID-19 vaccines. Many P/Ts recognized the important role public health will have in the development of communication strategies to counter these concerns. A Statistics Canada survey in June 2020 reported that 76.5% of Canadians would be very likely or somewhat likely to get a COVID-19 vaccine when available (5), but data from a national Leger survey in November 2020 estimate this number to be 65% (6).

P/Ts also highlighted the challenging logistics of vaccine delivery and the need to ensure that vaccination clinics follow public health restrictions. Multiple P/Ts viewed the 2020-21 seasonal influenza program as a trial for how COVID-19 vaccine delivery may occur. Following the H1N1 pandemic, it was noted that well-functioning influenza vaccination programs are essential for ensuring that adequate infrastructure is available for pandemic vaccination response (2). Guidance on strategies for influenza vaccine delivery during the pandemic were provided by NACI early in the pandemic (7).

Having a unified approach to COVID-19 vaccination in Canada may be beneficial for providing consistent public messaging, and clarifying why certain priority groups have been selected for early vaccination. Public communication strategies are important to prevent hesitancy and mistrust (2). Furthermore, a unified approach to vaccination may improve equity and produce cost-savings (8). Critics of Canada’s long-standing provincial and territorial variability in immunization programs and schedules have argued that lack of consistency in eligibility and modes of delivery results in inconsistencies in public messaging which can undermine public confidence when the rationale for differences is unclear. (8,9). Conversely, diversity across P/Ts enables flexibility to adapt to the unique circumstances of each jurisdiction, given the variation in geography, population, and COVID-19 cases across P/Ts. Although P/Ts will inevitably develop their own plans for COVID-19 vaccination, results from this study suggest that there will likely be many similarities.

### Strengths and Limitations

A strength of the current study was the wide variety of perspectives that were obtained on COVID-19 vaccination program planning from most P/Ts. As well, the use of key informant interviews in this study allowed us to gather in-depth perspectives on COVID-19 vaccination program planning in each P/T. However, as only a few select individuals were interviewed from each P/T, the perspectives gathered are not representative of entire P/Ts. Furthermore, there may be variation in individual perspectives across a single P/T, although the perspectives shared were very consistent within a given P/T. As well, interviews were conducted during a period when COVID-19 vaccination planning was in its early stages. It will be interesting to confirm whether early plans have changed since the release of NACI guidance documents (3,4).

### Implications

The implementation of COVID-19 vaccination programs in Canada is in the very early stages. There is an opportunity to expand on this study’s findings through a variety of research avenues, including the assessment of each P/T’s finalized COVID-19 vaccination plan, and how variation in vaccination programs ultimately affects vaccine uptake and effectiveness in each P/T. This study adds to existing literature by synthesizing P/T public health perspectives on COVID-19 vaccination programs. Results can inform policymakers and program planners and can assist NACI in future development of national guidelines. As well, we anticipate that the information in this study will enable P/Ts to learn from one another by comparing their approach to COVID-19 vaccination with others across Canada.

## Conclusion

The current study’s findings show that Canadian P/Ts are facing similar challenges in planning for COVID-19 vaccination. The majority will be relying on NACI recommendations regarding how to allocate limited vaccine supply. Further research is needed to evaluate the success of provincial/territorial COVID-19 vaccination programs, once they are implemented.

## Data Availability

Data are not publicly available, due to the potential for participants to be identifiable through the data.

## Authors’ Statement

SMacDonald – Conceptualization, methodology, funding acquisition, supervision, formal analysis, writing (original draft, and review and editing)

HS – Investigation, data curation, formal analysis, writing (original draft, and review and editing)

SW – Conceptualization, methodology, writing (review and editing)

SMeyer – Conceptualization, methodology, writing (review and editing)

AG – Conceptualization, methodology, funding acquisition, writing (review and editing)

AA – Investigation, data curation, supervision, project administration, formal analysis, writing (review and editing)

MS – Conceptualization, methodology, writing (review and editing)

## Conflict of Interest

SMacDonald is supported by a salary award from the Canadian Child Health Clinician Scientist Program. MS is supported via salary awards from the BC Children’s Hospital Foundation, the Canadian Child Health Clinician Scientist Program and the Michael Smith Foundation for Health Research. MS has been an investigator on projects funded by GlaxoSmithKline, Merck, Pfizer, Sanofi-Pasteur, Seqirus, Symvivo and VBI Vaccines. All funds have been paid to his institute, and he has not received any personal payments.

## Funding

This work was funded by the Canadian Institutes of Health Research.

## Contributors

This was part of a larger project conducted by the COVImm study team, which included the named authors, as well as: M Tunis; K Benzies; J Bettinger; M Driedger; E Dubé; R Humble; M Kiely; N MacDonald; E Rafferty; and J Robinson. We express particular thanks to M Kiely for her assistance with the interviews.

## Notes

### Author Declarations

Ethical approval for this study was obtained from the Health Research Ethics Board at the University of Alberta.

